# Predicting age from hearing test results with machine learning reveals the genetic and environmental factors underlying accelerated auditory aging

**DOI:** 10.1101/2021.07.05.21260048

**Authors:** Alan Le Goallec, Samuel Diai, Théo Vincent, Chirag J. Patel

**Author notes:** Co-first authors (Alan Le Goallec, Samuel Diai). **Contact information:** Chirag J Patel.

## Abstract

With the aging of the world population, age-related hearing loss (presbycusis) and other hearing disorders such as tinnitus become more prevalent, leading to reduced quality of life and social isolation. Unveiling the genetic and environmental factors leading to age-related auditory disorders could suggest lifestyle and therapeutic interventions to slow auditory aging. In the following, we built the first machine learning-based hearing age predictor by training models to predict chronological age from hearing test results (root mean squared error=7.10±0.07 years; R-Squared=31.4±0.8%). We defined hearing age as the prediction outputted by the model on unseen samples, and accelerated auditory aging as the difference between a participant’s hearing age and age. We then performed a genome wide association study [GWAS] and found that accelerated hearing aging is 14.1±0.4% GWAS-heritable. Specifically, accelerated auditory aging is associated with 662 single nucleotide polymorphisms in 243 genes (e.g *OR2B4P*, involved in smell perception). Similarly, it is associated with biomarkers (e.g cognitive tests), clinical phenotypes (e.g chest pain), diseases (e.g depression), environmental (e.g smoking, sleep) and socioeconomic (e.g income, education, social support) variables. The hearing age predictor could be used to evaluate the efficiency of emerging rejuvenation therapies on hearing.

## Introduction

With age, the auditory system undergoes important changes ^1^, leading to the development of age related hearing disorders such as presbycusis (age-related hearing loss) ^2^. Hearing loss is associated with social isolation ^3^, depression ^4^, cognitive decline ^5^, dementia ^6^. Age-related hearing loss affects one third of the population after age 65 ^7^ and with the world population aging ^8^, the burden of this disease is projected to increase ^9^.

To study the aging process of a specific organ, biological age predictors can be built by training machine learning algorithms to predict age (also referred as “chronological age”) from biomedical datasets related to the organ’s health. The prediction outputted by the model on unseen samples can be interpreted as the participant’s organ age. For example, heart age predictors have been trained on magnetic resonance images ^10^ [MRIs] and electrocardiograms ^11^, and brain age has been predicted from brain MRIs ^12^. Although the relationship between hearing loss and age has been clearly established ^1^, no hearing age predictor has been developed, to our knowledge.

In the following, we build the first hearing age predictor by training machine learning algorithms to predict age from 229,410 hearing tests collected from 37-82 year-old UK Biobank [UKB] ^13^ participants. We defined hearing age as the prediction outputted by the model on unseen samples, and accelerated auditory aging as the difference between hearing age and age. For example, a 60 year-old participant predicted to be 70 years old by the model has a hearing age of 70 years and is an accelerated auditory ager of ten years. We then performed a genome wide association study [GWAS] to estimate the heritability of accelerated auditory aging and to identify single nucleotide polymorphisms [SNPs] associated with this phenotype. Similarly, we performed an X-Wide Association Study [XWAS] to identify biomarkers, clinical phenotypes, diseases, environmental and socioeconomic variables associated with accelerated auditory aging. (Figure 1)

**Figure 1:**
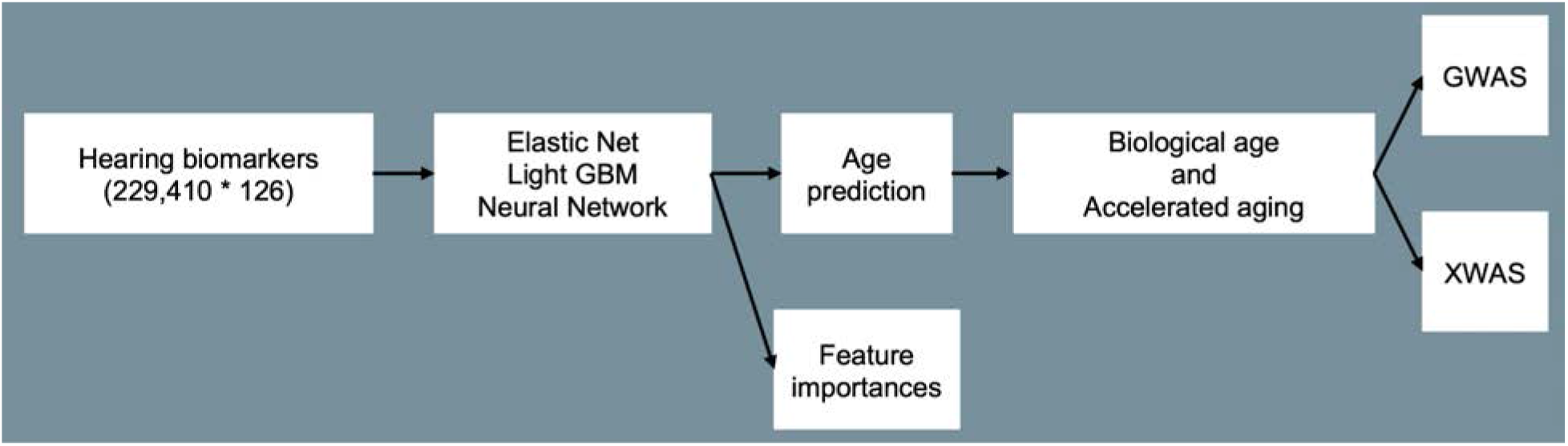
Overview of the analytic pipeline.

## Results

### Hearing tests explain 31% of the variance in age

We predicted age from 229,410 hearing test results (126 features for each sample) using an ensemble of an elastic net, a gradient boosted machine [GBM] and a shallow, fully connected neural network with a R-Squared [R^2^] of 48.6±0.4% and a root mean squared error of 5.92±0.02 years. The best performing non-ensemble model was the GBM, and both non-linear models (GBM, neural network) outperformed the linear model (elastic net). (Table 1)

**Table 1:** Age prediction performance.

| Model | R-Squared (%) | RMSE (years) |
| --- | --- | --- |
| Ensemble | $31.4 \pm 0.8\%$ | $7.10 \pm 0.07$ |
| Elastic Net | $25.5 \pm 0.7\%$ | $7.40 \pm 0.06$ |
| GBM | $31.1 \pm 0.9\%$ | $7.11 \pm 0.07$ |
| Neural Network | $31.0 \pm 0.8\%$ | $7.12 \pm 0.07$ |

We found that the most important scalar features are the duration of the hearing test and the time to press next for the first tests, which all had a positive coefficient in the elastic net (R^2^=25.5±0.7%). The complete feature importances are found in Table S1.

### Genetic factors and heritability of accelerated auditory aging

We performed a genome wide association study [GWAS]. Accelerated auditory aging is 14.1±0.4% GWAS-heritable and 662 single nucleotide polymorphisms [SNPs] in 243 genes are significantly associated with this phenotype. The six peaks highlighted by the GWAS are (1) *OR2B4P* (Olfactory Receptor Family 2 Subfamily B Member 4 Pseudogene, a GPCR involved in smell perception); (2) *DDR1* (Discoidin Domain Receptor Tyrosine Kinase 1, a cell surface receptor); (3) *APOC1* (Apolipoprotein C1, involved in high density lipoprotein (HDL) and very low density lipoprotein (VLDL) metabolism); (4) *TSNARE1* (T-SNARE Domain Containing 1, involved in vesicle fusion and linked to schizophrenia); (5) *AKAP6* (A-Kinase Anchoring Protein 6); and (6) *OPCML* (Opioid Binding Protein/Cell Adhesion Molecule Like, possibly involved in opioid receptor function). (Figure 2)

**Figure 2:**
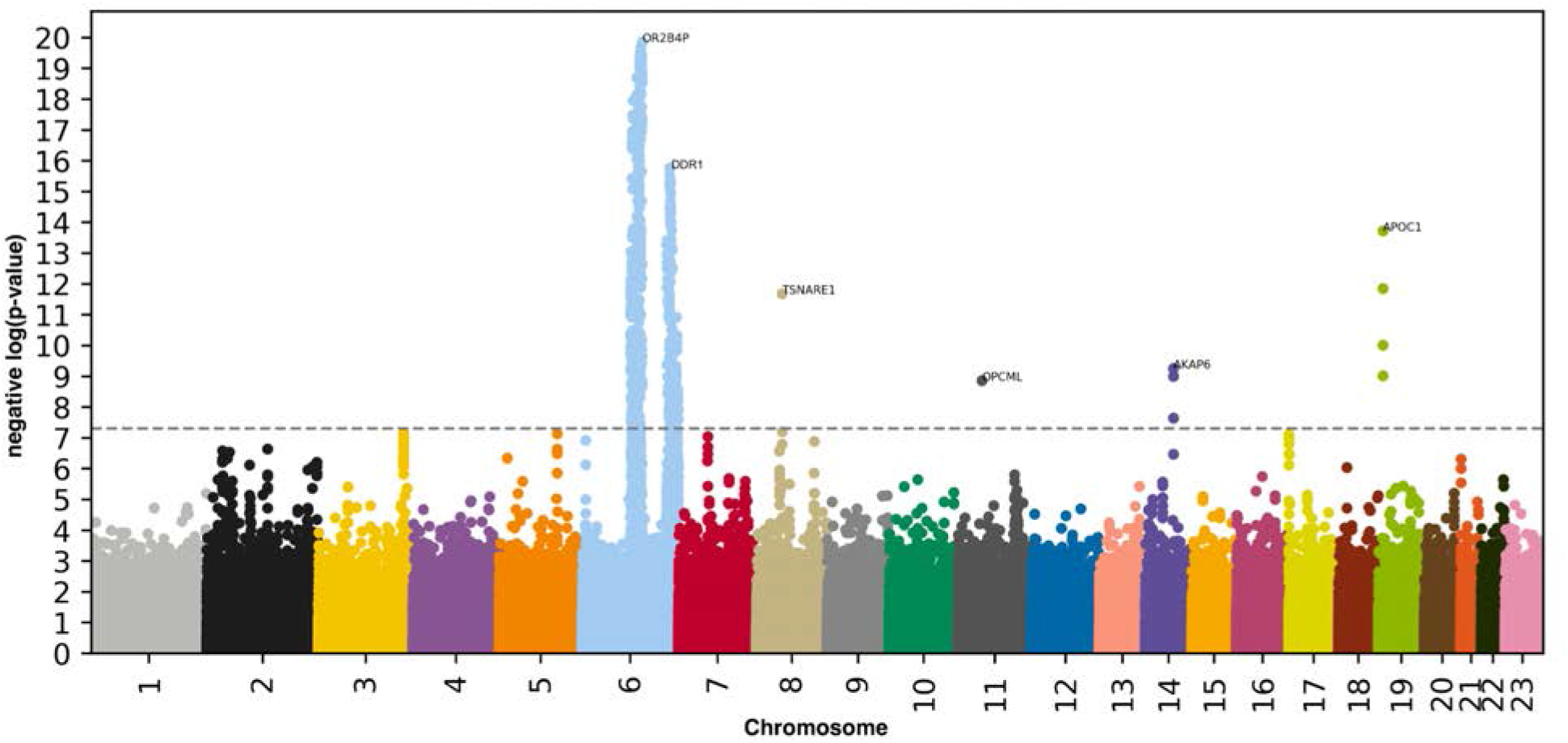
GWAS results - SNPs associated with accelerated auditory aging. -log10(p-value) vs. chromosomal position of locus. Dotted line denotes 5×10^−8^.

### Biomarkers, clinical phenotypes, diseases, environmental and socioeconomic variables associated with accelerated auditory aging

We use “X” to refer to all nongenetic variables measured in the UK Biobank (biomarkers, clinical phenotypes, diseases, family history, environmental and socioeconomic variables). We performed an X-Wide Association Study [XWAS] to identify which of the 4,372 biomarkers classified in 21 subcategories (Table S3), 187 clinical phenotypes classified in 11 subcategories (Table S6), 2,073 diseases classified in 26 subcategories (Table S9), 92 family history variables (Table S12), 265 environmental variables classified in nine categories (Table S13), and 91 socioeconomic variables classified in five categories (Table S16) are associated (p-value threshold of 0.05 and Bonferroni correction) with accelerated auditory aging. We summarize our findings in Table S4, Table S5, Table S7, Table S8, Table S10, Table S11, Table S14, Table S15, Table S17, Table S18, where we list the three variables of each X-subcategory (e.g spirometry biomarkers) most associated with each accelerated aging dimension. The full results can be exhaustively explored at https://www.multidimensionality-of-aging.net/xwas/univariate_associations.

#### Biomarkers associated with accelerated auditory aging

The three biomarker categories most associated with accelerated auditory aging are body impedance, prospective memory (a cognitive test) and trail making (another cognitive test) (Table S4). Specifically, 100.0% of body impedance biomarkers are associated with accelerated auditory aging, with the three largest associations being with leg left impedance (correlation=.025), right leg impedance (correlation=.024), and right arm impedance (correlation=.023). 78.6% of prospective memory tests are associated with accelerated auditory aging, with the three largest associations being with giving a wrong first answer (correlation=.126), number of attempts (correlation=.100), and time to answer (correlation=.087). 75.0% of trail making biomarkers are associated with accelerated auditory aging, with the three largest associations being with duration to complete the alphanumeric path #2 (correlation=.129), duration to complete the alphanumeric path #1 (correlation=.113), and total errors traversing the alphanumeric path #2 (correlation=.076). Hearing came in fifth position (69.0% of biomarkers associated with accelerated aging), with the three variables most associated being speech-reception-threshold estimate for the right ear (correlation=.396), mean signal-to-noise ratio for the right ear (correlation=.396), and speech-reception-threshold estimate for the left ear (correlation=.396).

Conversely, the three biomarker categories most associated with decelerated auditory aging are anthropometry, hand grip strength and symbol digit substitution (a cognitive test) (Table S5). Specifically, 100.0% of anthropometry biomarkers are associated with decelerated auditory aging, with the three largest associations being with seated height (correlation=.082), sitting height (correlation=.070), and standing height (correlation=.068). 100.0% of hand grip strength biomarkers are associated with decelerated auditory aging, with the two associations being with right hand grip strength (correlation=.052) and left hand grip strength (correlation=.042). 100.0% of symbol digit substitution biomarkers are associated with decelerated auditory aging, with the two associations being with number of symbol digit matches attempted (correlation=.160) and number of symbol digit matches made correctly (correlation=.155). Hearing came in 25th position (4.8% of biomarkers associated with decelerated auditory aging), with the three largest associations being with first triplet correct for the left ear (correlation=.114), first triplet correct for the right ear (correlation=.111) and second triplet correct for the left ear (correlation=.087).

#### Clinical phenotypes associated with accelerated auditory aging

The three clinical phenotype categories most associated with accelerated auditory aging are chest pain, breathing and claudication (Table S7). Specifically, 100.0% of chest pain phenotypes are associated with accelerated auditory aging, with the three largest associations being with chest pain or discomfort walking normally (correlation=.044), chest pain due to walking ceases when standing still (correlation=.029), and chest pain or discomfort (correlation=.028). 100.0% of breathing phenotypes are associated with accelerated auditory aging, with the two associations being with shortness of breath walking on level ground (correlation=.049) and wheeze or whistling in the chest in the last year (correlation=.027). 76.9% of claudication phenotypes are associated with accelerated auditory aging, with the three largest associations being with leg pain in calf/calves (correlation=.049), leg pain when walking uphill or hurrying (correlation=.046), and leg pain when standing still or sitting (correlation=.045). Hearing phenotypes came in fifth position (64.3% of phenotypes associated with accelerated auditory aging), with the three largest associations being with noisy workplace (correlation=.080), hearing aid user (correlation=.077), and hearing difficulty/problems (correlation=.075).

Conversely, the three clinical phenotype categories most associated with decelerated auditory aging are cancer screening (one association: ever had prostate specific antigen test; correlation=.029), oral health (one association: no bucal/dental problem; correlation=.031) and general health (one association: weight loss during the last year; correlation=.017). One hearing variable was associated with decelerated auditory aging: never experienced tinnitus (correlation=.038) (Table S8).

#### Diseases associated with accelerated auditory aging

The three disease categories most associated with accelerated auditory aging are mental disorders, a category encompassing diverse diseases, and vascular diseases (Table S10). Specifically, 32.4% of mental diseases are associated with accelerated auditory aging, with the three largest associations being with mental and behavioral disorders due to use of alcohol (correlation=.036), depressive episode (correlation=.034), and mental and behavioral disorders due to use of tobacco (correlation=.032). 30.7% of the diseases in the diverse category are associated with accelerated auditory aging, with the three largest associations being with symptoms and signs involving cognitive functions and awareness (correlation=.026), symptoms and signs involving the nervous and musculoskeletal systems (correlation=.025), and abnormalities of gait and mobility (correlation=.020). 19.5% of vascular diseases are associated with accelerated auditory aging, with the three largest associations being with sequelae of cerebrovascular disease (correlation=.024), varicose veins of lower extremities (correlation=.021), and cerebrovascular diseases (correlation=.018). We found no disease associated with decelerated auditory aging (Table S11).

#### Environmental variables associated with accelerated auditory aging

The three environmental variable categories most associated with accelerated auditory aging are early life factors, smoking and sleep (Table S14). Specifically, 56.2% of early life factors variables are associated with accelerated auditory aging, with the three largest associations being with countries of birth (not in the UK, Wales, and Republic of Ireland; respective correlations of .035, .029 and .025). 45.8% of smoking variables are associated with accelerated auditory aging, with the three largest associations being with age when started smoking (correlation=.041), difficulty not smoking for one day (correlation=.039), and number of cigarettes smoked daily (correlation=.038). 42.9% of sleep variables are associated with accelerated auditory aging, with the three largest associations being with daytime dozing/sleeping/narcolepsy (correlation=.045), nap during the day (correlation=.038), and snoring (correlation=.017).

Conversely, the three environmental variable categories most associated with decelerated auditory aging are electronic devices, physical activity and smoking (Table S15). Specifically, 50.0% of electronic devices variables are associated with decelerated auditory aging, with the three largest associations included playing computer video games (correlation=.097), length of mobile phone use (correlation=.059), and hands-free device/speakerphone use with mobile phone in the last three months (correlation=.035). 45.7% of physical activity variables are associated with decelerated auditory aging, with the three largest associations being with driving faster than motorway speed limit (correlation=.118), time spent using the computer (correlation=.101), and types of transport used: car/motor vehicle (correlation=.070). 25.0% of smoking environmental variables are associated with decelerated auditory aging, with the three largest associations being with time waking to the first cigarette (correlation=.041), smoking status: previous (correlation=.040), and past tobacco smoking: smoked on most or all days (correlation=.038).

#### Socioeconomic variables associated with accelerated auditory aging

The three socioeconomic variable categories most associated with accelerated auditory aging are sociodemographics, education and household (Table S17). Specifically, 57.1% of sociodemographic variables are associated with accelerated auditory aging, with the three largest associations being with private healthcare (correlation=.057), receiving attendance allowance (correlation=.030), and attendance/disability/mobility: prefer not to answer (correlation=.025). 37.5% of education variables are associated with accelerated auditory aging, with the three largest associations being with not having a qualification among the ones listed (correlation=.117), qualification: GSEs or equivalent (correlation=.052), and qualifications: prefer not to answer (correlation=.032). 36.4% of household variables are associated with accelerated auditory aging, with the three largest associations being with renting from local authority, local council or housing association (correlation=.080), living in a flat, a maisonette or an apartment (correlation=.056), and renting from a private landlord or letting agency (correlation=.031).

Conversely, the three socioeconomic variable categories most associated with decelerated auditory aging are education, employment and social support (Table S18). Specifically, 37.5% of education variables are associated with decelerated auditory aging, with the three largest associations being with having a college or university degree (correlation=.077), A/AS levels or equivalent (correlation=.047), and O levels/GCSEs or equivalent (correlation=.026). 34.8% of employment socioeconomic variables are associated with decelerated auditory aging, with the three largest associations being with length of working week for main job (correlation=.083), being in paid employment or self-employed (correlation=.078), and being retired (correlation=.057). 22.2% of social support variables are associated with decelerated auditory aging, with the two associations being with leisure/social activities: sports club or gym (correlation=.035) and being able to confide (correlation=.016).

### Predicting accelerated aging from biomarkers, clinical phenotypes, diseases, environmental variables and socioeconomic variables

We predicted accelerated auditory aging using variables from the different X-datasets categories (biomarkers, clinical phenotypes, diseases, environmental variables and socioeconomic variables). Specifically we built a model using the variables from each of their respective subcategories (e.g blood pressure biomarkers), and found that no dataset could explain more than 5% of the variance in accelerated auditory aging.

## Discussion

We built the first hearing age predictor by training machine learning algorithms to predict age from hearing test results (RMSE=7.10±0.07 years; R^2^=31.4±0.8%). Non-linear models significantly outperformed the linear model (R^2^=31.1±0.9% vs. 25.5±0.7%), suggesting that information regarding aging is encoded non-linearly or in complex interactions between the 126 hearing test-derived predictors we used to train the models.

Accelerated auditory aging is 14.1±0.4% GWAS-heritable and is associated with SNPs in genes whose function is not directly connected to hearing (e.g *OR2B4P*, involved in olfaction, *APOC1*, involved in lipoprotein metabolism, *TSNARE1*, linked to schizophrenia, *OPCML*, involved in opioid receptor function). Similarly, accelerated auditory aging is associated with biomarkers, clinical phenotypes and diseases in diverse organ systems such as blood and urine biochemistry, blood count, eyesight, cardiovascular health, lung function, musculoskeletal function, oral health, metabolic disease and others. Interestingly, auditory aging is also associated with facial aging. These observations suggest that auditory aging is connected to a general aging process affecting all organs. In particular, accelerated auditory aging is associated with brain, cognitive and mental health (e.g cognitive tests, brain MRI features and volumes, mental disorders such as depression), which is coherent with the literature on the effect of age-related hearing loss on the brain and cognitive function ^5^. Similarly, a clear link has been established between cardiovascular health and hearing function ^14^. We explore the connection between hearing aging and aging in other organ systems more in depth in a different paper ^15^.

In terms of environmental factors, we found that accelerated auditory aging is associated with working in a noisy environment, smoking and poor sleep, as reported in the literature ^16–18^. In contrast, some of our findings run contrary to the literature. For example, we found that accelerated auditory aging is associated with physical activity (e.g frequency and duration of moderate activity, duration of walks) ^19^, and that decelerated auditory aging is associated with driving faster that the motorway speed limit, commuting using a motor vehicle ^20^, time spent using a computer or playing video games ^21^, length of mobile phone use and use of hands-free devices ^22^. We hypothesize that these associations might be confounded by unaccounted factors (we only corrected the partial correlations for age, sex and ethnicity). For alcohol intake, diet and medications, the associations are product specific. Alcohol intake frequency, bread intake and tea intake are associated with accelerated aging, whereas drinking red wine, ground coffee and skimmed milk, eating wholemeal/wholegrain bread and taking glucosamine (a supplement for cartilage health) and ibuprofen are associated with decelerated aging. Conflicting associations between alcohol intake and hearing loss have been reported in the literature ^23–26^. Others have reported associations between diet and hearing function ^27^.

In terms of socioeconomic status, wealth and education are associated with decelerated auditory aging. Possible explanations include that wealthier individuals are less likely to be exposed to noise pollution, and that wealth is associated with lower aging rates overall, with the richest 1% US males/females respectively living 14.6±0.2/10.1±0.2 years longer than their poorest 1% counterparts ^28^. Low social support (no leisure/social activities, no one to confide in) are associated with accelerated auditory aging, as reported in the literature ^29^.

A limitation of our work is that UKB is an observational, cross-sectional study. As a consequence, the correlation that we report between accelerated auditory aging and biomarkers, clinical phenotypes, diseases, environmental and socioeconomic variables do not allow us to infer causation. Another notable limitation of our work is the narrow age range covered by UKB (37-82 years), which prevented us from studying age-related hearing loss in younger and older participants. Hearing loss is, for example, a particularly prevalent condition for centenarians ^30^.

## Methods

### Data and materials availability

We used the UK Biobank (project ID: 52887). The code can be found at https://github.com/Deep-Learning-and-Aging. The results can be interactively and extensively explored at https://www.multidimensionality-of-aging.net/. We will make the hearing age phenotype available through UK Biobank upon publication. The GWAS results can be found at https://www.dropbox.com/s/59e9ojl3wu8qie9/Multidimensionality_of_aging-GWAS_results.zip?dl=0.

### Software

Our code can be found at https://github.com/Deep-Learning-and-Aging. For the genetics analysis, we used the BOLT-LMM ^31,32^ and BOLT-REML ^33^ softwares. We coded the parallel submission of the jobs in Bash ^34^.

### Cohort Dataset: Participants of the UK Biobank

We leveraged the UK Biobank^13^ cohort (project ID: 52887). The UKB cohort consists of data originating from a large biobank collected from 502,211 de-identified participants in the United Kingdom that were aged between 37 years and 74 years at enrollment (starting in 2006). The dataset includes 229,410 hearing test samples, with 126 measurements collected for each. The Harvard internal review board (IRB) deemed the research as non-human subjects research (IRB: IRB16-2145).

### Data types and Preprocessing

The data preprocessing step is different for demographic variables and hearing test biomarkers. For demographics variables, first, we removed out the UKB samples for which age or sex was missing. For sex, we used the genetic sex when available, and the self-reported sex when genetic sex was not available. We computed age as the difference between the date when the participant attended the assessment center and the year and month of birth of the participant to estimate the participant’s age with greater precision. We one-hot encoded ethnicity. For the biomarkers, we did not perform any preprocessing, aside from the normalization that is described under cross-validation further below. The complete list of hearing test biomarkers can be found in Table S3 under “Hearing”.

### Machine learning algorithms

We used three different algorithms to predict age. Elastic Nets [EN] (a regularized linear regression that represents a compromise between ridge regularization and LASSO regularization), Gradient Boosted Machines [GBM] (LightGBM implementation ^35^), and Neural Networks [NN]. The choice of these three algorithms represents a compromise between interpretability and performance. Linear regressions and their regularized forms (LASSO ^36^, ridge ^37^, elastic net ^38^) are highly interpretable using the regression coefficients but are poorly suited to leverage non-linear relationships or interactions between the features and therefore tend to underperform compared to the other algorithms. In contrast, neural networks ^39,40^ are complex models, which are designed to capture non-linear relationships and interactions between the variables. However, tools to interpret them are limited ^41^ so they are closer to a “black box”. Tree-based methods such as random forests ^42^, gradient boosted machines ^43^ or XGBoost ^44^ represent a compromise between linear regressions and neural networks in terms of interpretability. They tend to perform similarly to neural networks when limited data is available, and the feature importances can still be used to identify which predictors played an important role in generating the predictions. However, unlike linear regression, feature importances are always non-negative values, so one cannot interpret whether a predictor is associated with older or younger age. We also performed preliminary analyses with other tree-based algorithms, such as random forests ^42^, vanilla gradient boosted machines ^43^ and XGBoost ^44^. We found that they performed similarly to LightGBM, so we only used this last algorithm as a representative for tree-based algorithms in our final calculations.

### Training, tuning and predictions

We split the entire dataset into ten data folds and performed a nested-cross validation. We describe the splitting of the data into different folds and the tuning procedures in greater detail in the Supplementary.

### Interpretability of the machine learning predictions

To interpret the models, we used the regression coefficients for the elastic nets, the feature importances for the GBMs, and a permutation test for the fully connected neural networks (Supplementary Methods).

### Ensembling to improve prediction and define aging dimensions

We ensembled the elastic net, the GBM and the neural network. We built each ensemble model separately on each of the ten data folds. For example, to build the ensemble model on the testing predictions of the data fold #1, we trained and tuned an elastic net on the validation predictions from the data fold #0 using a 10-folds inner cross-validation, as the validation predictions on fold #0 and the testing predictions on fold #1 are generated by the same model. We used the same hyperparameters space and Bayesian hyperparameters optimization method as we did for the inner cross-validation we performed during the tuning of the non-ensemble models.

To summarize, the testing ensemble predictions are computed by concatenating the testing predictions generated by ten different elastic nets, each of which was trained and tuned using a 10-folds inner cross-validation on one validation data fold (10% of the full dataset) and tested on one testing fold. This is different from the inner-cross validation performed when training the non-ensemble models, which was performed on the “training+validation” data folds, so on 9 data folds (90% of the dataset).

### Evaluating the performance of models

We evaluated the performance of the models using two different metrics: R-Squared [R^2^] and root mean squared error [RMSE]. We computed a confidence interval on the performance metrics in two different ways. First, we computed the standard deviation between the different data folds. The test predictions on each of the ten data folds are generated by ten different models, so this measure of standard deviation captures both model variability and the variability in prediction accuracy between samples. Second, we computed the standard deviation by bootstrapping the computation of the performance metrics 1,000 times. This second measure of variation does not capture model variability but evaluates the variance in the prediction accuracy between samples.

### Hearing age definition

We defined the hearing age (also called biological age) of participants as the prediction generated by the model, after correcting for the bias in the residuals. We indeed observed a bias in the residuals. For each model, participants on the older end of the chronological age distribution tend to be predicted younger than they are. Symmetrically, participants on the younger end of the chronological age distribution tend to be predicted older than they are. This bias does not seem to be biologically driven. Rather it seems to be statistically driven, as the same 60-year-old individual will tend to be predicted younger in a cohort with an age range of 60-80 years, and to be predicted older in a cohort with an age range of 60-80. We ran a linear regression on the residuals as a function of age for each model and used it to correct each prediction for this statistical bias.

After defining biological age as the corrected prediction, we defined accelerated aging as the corrected residuals. For example, a 60-year-old whose hearing test results predicted an age of 70 years old after correction for the bias in the residuals is estimated to have a hearing age of 70 years, and an accelerated auditory aging of ten years.

It is important to understand that this step of correction of the predictions and the residuals takes place after the evaluation of the performance of the models but precedes the analysis of the biological ages properties.

### Genome-wide association study of accelerated aging

The UKB contains genome-wide genetic data for 488,251 of the 502,492 participants^45^ under the hg19/GRCh37 build. We used the average accelerated aging value over the different samples collected over time (see Supplementary - Generating average predictions for each participant). Next, we performed genome wide association studies [GWASs] to identify single-nucleotide polymorphisms [SNPs] associated with accelerated auditory aging using BOLT-LMM ^31,32^ and estimated the SNP-based heritability for this phenotype. We used the v3 imputed genetic data to increase the power of the GWAS, and we corrected all of them for the following covariates: age, sex, ethnicity, the assessment center that the participant attended when their DNA was collected, and the 20 genetic principal components precomputed by the UKB. We used the linkage disequilibrium [LD] scores from the 1,000 Human Genomes Project ^46^. To avoid population stratification, we performed our GWAS on individuals with White ethnicity.

#### Identification of SNPs associated with accelerated aging

We identified the SNPs associated with accelerated aging dimensions using the BOLT-LMM ^31,32^ software (p-value of 5e-8). The sample size for the genotyping of the X chromosome is one thousand samples smaller than for the autosomal chromosomes. We therefore performed two GWASs for each aging dimension. (1) excluding the X chromosome, to leverage the full autosomal sample size when identifying the SNPs on the autosome. (2) including the X chromosome, to identify the SNPs on this sex chromosome. We then concatenated the results from the two GWASs to cover the entire genome, at the exception of the Y chromosome. We plotted the results using a Manhattan plot and a volcano plot. We used the bioinfokit ^47^ python package to generate the Manhattan plots. We generated quantile-quantile plots [Q-Q plots] to estimate the p-value inflation as well.

#### Heritability and genetic correlation

We estimated the heritability of the accelerated aging dimensions using the BOLT-REML ^33^ software. We included the X chromosome in the analysis and corrected for the same covariates as we did for the GWAS. Using the same software and parameters, we computed the genetic correlations between accelerated aging in the different aging dimensions. We annotated the significant SNPs with their matching genes using the following four steps pipeline. (1) We annotated the SNPs based on the rs number using SNPnexus ^48–52^. When the SNP was between two genes, we annotated it with the nearest gene. (2) We used SNPnexus to annotate the SNPs that did not match during the first step, this time using their genomic coordinates. After these two first steps, 30 out of the 9,697 significant SNPs did not find a match. (3) We annotated these SNPs using LocusZoom ^53^. Unlike SNPnexus, LocusZoom does not provide the gene types, so we filled this information with GeneCards ^54^. After this third step, four genes were not matched. (4) We used RCSB Protein Data Bank ^55^ to annotate three of the four missing genes.

### Non-genetic correlates of accelerated aging

We identified non-genetically measured (i.e factors not measured on a GWAS array) correlates of accelerated auditory aging, which we classified in six categories: biomarkers, clinical phenotypes, diseases, family history, environmental, and socioeconomic variables. We refer to the union of these association analyses as an X-Wide Association Study [XWAS]. (1) We define as biomarkers the scalar variables measured on the participant, which we initially leveraged to predict age (e.g. blood pressure, Table S3). (2) We define clinical phenotypes as other biological factors not directly measured on the participant, but instead collected by the questionnaire, and which we did not use to predict chronological age. For example, one of the clinical phenotypes categories is eyesight, which contains variables such as “wears glasses or contact lenses”, which is different from the direct refractive error measurements performed on the participants, which are considered “biomarkers” (Table S6). (3) Diseases include the different medical diagnoses categories listed by UKB (Table S9). (4) Family history variables include illnesses of family members (Table S12). (5) Environmental variables include alcohol, diet, electronic devices, medication, sun exposure, early life factors, medication, sun exposure, sleep, smoking, and physical activity variables collected from the questionnaire (Table S13). (6) Socioeconomic variables include education, employment, household, social support and other sociodemographics (Table S16). We provide information about the preprocessing of the XWAS in the Supplementary Methods.

## Data Availability

https://www.multidimensionality-of-aging.net/

https://www.dropbox.com/s/59e9ojl3wu8qie9/Multidimensionality_of_aging-GWAS_results.zip?dl=0

## Author Contributions

**Alan Le Goallec:** (1) Designed the project. (2) Supervised the project. (3) Ensembled the models, evaluated their performance and computed auditory age. (4) Performed the genome wide association study. (5) Designed the website. (6) Wrote the manuscript.

**Samuel Diai:** (1) Predicted chronological age from the hearing test. (2) Coded the algorithm to obtain balanced data folds across the different datasets. (3) Wrote the python class to build an ensemble model using a cross-validated elastic net. (4) Performed the X-wide association study. (5) Implemented a first version of the website https://www.multidimensionality-of-aging.net/.

**Théo Vincent:** (1) Website data engineer. (2) Implemented a second version of the website https://www.multidimensionality-of-aging.net/.

**Chirag J. Patel:** (1) Supervised the project. (2) Edited the manuscript. (3) Provided funding.

## Acknowledgments

We would like to thank Raffaele Potami from Harvard Medical School research computing group for helping us utilize O2’s computing resources. We thank HMS RC for computing support. We also want to acknowledge UK Biobank for providing us with access to the data they collected. The UK Biobank project number is 52887.

## Funding

NIEHS R00 ES023504

NIEHS R21 ES25052.

NIAID R01 AI127250

NSF 163870

MassCATS, Massachusetts Life Science Center

Sanofi

The funders had no role in the study design or drafting of the manuscript(s).

