## Supplementary Information for "Predicting age from hearing test results with machine learning reveals the genetic and environmental factors underlying accelerated auditory aging"

### Supplementary Methods, Figures, Tables and References

#### Table of contents

**Table of contents** 1

**Methods** 2
- Hardware 2
- Software 2
- Training, tuning and predictions 2
  - Data splitting 2
  - Nested cross-validation 3
  - Bayesian hyperparameters optimization 3
  - Example 4
- Generating average predictions for each participant 6
- Interpretability of the predictions 8
- Non-genetic correlates of accelerated aging 8
  - Imputation of the non-genetic X-variables 9
  - X-Wide Association Studies 10
  - Prediction of accelerated aging 11

**Supplementary Figures** 12

**Supplementary Tables** 13

**Supplementary References** 18

### Methods

#### Hardware

We performed the computation for this project on Harvard Medical School's compute cluster, with access to both central processing units [CPUs] and general processing units [GPUs] (Tesla-M40, Tesla-K80, Tesla-V100) via a Simple Linux Utility for Resource Management [SLURM] scheduler.

#### Software

We coded the project in Python [1] and used the following libraries: NumPy [2,3], Pandas [4], Matplotlib [5], Plotly [6], Python Imaging Library [7], SciPy [8–10], Scikit-learn [11], LightGBM [12], XGBoost [13], Hyperopt [14], TensorFlow 2 [15], Keras [16], Keras-vis [17], iNNvestigate [18]. We used Dash [19] to code the website on which we shared the results. We set the seed for the os library, the numpy library, the random library and the tensorflow library to zero.

#### Training, tuning and predictions

##### Data splitting

We split the 676,787 samples into ten data folds, while keeping all samples from the same participant in the same fold. To ensure this, we split the 502,211 participants' ids (referred to by UKB as "eid") into ten different buckets of the same size.

#### Nested cross-validation

Cross-validation is a method to tune the regularization of models and prevent overfitting [20]. For the models inputting scalar data (Figure 1A in green), we tuned the hyperparameters and generated a testing prediction for each sample using a nested 10x9-folds cross-validation. We refer to the two nested cross-validations as the "outer" and the "inner" cross-validations. The outer-cross validation is used to generate an unbiased testing prediction for each sample, as opposed to a simple split of the data into a "training+validation" set on one hand, and a testing set on the other hand, which would only generate a testing prediction for one tenth of the dataset. The inner cross-validation is used to tune the hyperparameters more precisely, leveraging the full inner cross-validation dataset as a validation set, as opposed to a simple data split of the "training+validation" dataset into a training and a validation sets, which would only use one data fold as the validation set to estimate the performance associated with a specific combination of hyperparameters. The nested cross-validation is illustrated in Table S20.

#### Bayesian hyperparameters optimization

To tune the hyperparameters, we used the Tree-structured Parzen Estimator Approach [21] [TPE] of the hyperopt python package [22]. TPE is a sequential Bayesian hyperparameters optimization method that iteratively suggests the next most promising hyperparameters combination as a function of the hyperparameters combinations that have already been tested, by building a probabilistic representation of the objective function. We set the number of iterations to 30. For each model, 30 different hyperparameter combinations are iteratively tested before selecting the best performing one. The hyperparameters names and their ranges defining the hyperparameters space can be found in Table S19. It might be of interest to other researchers that we initially tuned the hyperparameters using a random search [23] with the same number of iterations, and we did not observe a significant improvement in the model's performance after implementing the Bayesian hyperparameters optimization.

#### Example

For the sake of clarity, let us walk through a concrete example, which is illustrated in Table S20. Suppose we want to generate unbiased predictions for every sample in a dataset using an elastic net. First, let us generate the testing prediction for the data fold F9, which is performed by the first fold of the outer cross-validation (outer cross-validation fold 0). We select the data fold F9 out of the ten data folds as the testing fold, and we select the remaining nine data folds as "training+validation" folds for the inner cross-validation. We scale and center the target (age) and the predictors using the mean and standard deviation values of the variables on the "training+validation" dataset. We then enter the first inner-cross validation.

For the first inner cross-validation fold, we select the data fold F8 as the validation set, and the remaining eight "training+validation" data folds as the training set. We re-scale and center age and the predictors in the training and the validation sets using the mean and standard deviation values of the training set. We train the model on the eight training data folds with the first hyperparameters combination sampled by the TPE algorithm (one value for alpha and one value for l1_ratio) and generate validation predictions on the validation fold (data fold F8), which we unscale. This completes the first of the nine inner cross-validation folds (Inner CV fold 0). We then permute the nine inner data folds. We scale the age and the predictors using the mean and standard deviation computed on the new training set. Then we train the model with the same first combination of hyperparameters on eight data folds, leaving aside the data fold F9 (still being used as the testing set for the outer cross-validation) and the data fold F7 (now being used as the validation set for the inner cross-validation). We then use the new trained model to generate validation predictions on the data fold F7, which we unscale. This completes the second of the nine inner-cross validation folds (Inner CV fold 1). We then reiterate these inner permutation and training processes seven more times, until every data fold in the nine “training+validation” data folds is used as the validation set once. At this point, we concatenate the validation predictions from these nine validation folds to obtain the overall validation predictions associated with the first hyperparameters combination, and compute the associated performance metric (e.g. RMSE). This completes the inner-cross validation for the first hyperparameters combination.

We then perform the same 9-folds inner cross-validation, this time with the second hyperparameters combination suggested by the TPE algorithm. We iterate this process 28 more times, until 30 different hyperparameters combinations have iteratively been tested. Next, we select the hyperparameter combination that yielded the best validation performance (e.g. minimum RMSE), and we retrain a model on the whole nine “training+validation” data folds (all data folds except for data fold #1), using this best performing hyperparameters combination. This completes the first inner cross-validation.

We then use the model to generate unbiased predictions on the unseen testing set (data fold F9) and record these predictions. By anticipation for the ensembling algorithm (see Methods - Models ensembling) we also need to compute validation predictions on the data fold F8. We do this by training a model on all the data folds aside from the validation fold (data fold F8) and the testing fold (data fold F9), with the selected hyperparameters combination. We then use this trained model to compute predictions on the validation fold (data fold F8) and record these predictions, after unscaling them. This completes the first of the ten outer cross-validation folds (outer cross-validation 0).

We then complete the second outer cross-validation fold (outer cross-validation 1), this time using the data fold F8 as the testing dataset, to obtain unbiased testing predictions on this data fold, as well as validation predictions on the data fold F7. We reiterate the process eight more times to obtain the testing and validation predictions on the remaining data folds. We then concatenate the testing predictions from the ten data folds to obtain our final testing predictions for the model. Similarly, we concatenate the validation predictions from the ten data folds to obtain our final testing predictions for the model, which will later be used during ensemble models building and model selection (see Methods - Models ensembling).

The final validation and testing predictions for each data fold are therefore not necessarily associated with the same hyperparameters combination. It is also important to notice that we performed a single outer cross-validation, but that we performed a separate inner-cross validation for each outer cross-validation fold (hence the word “nested”), for a total of ten inner cross-validations per outer cross-validation fold.

#### Generating average predictions for each participant

We generated an average prediction for each individual, reported to UKB’s instance 0. We walk through an example. Let us assume a participant had two glucose levels samples collected from them in instances 2 and 3, respectively at age 70 and 80. Let us assume that the age predictions were respectively 64 and 78, so the residuals are respectively -6 years and -2 years, for an average of -4 years. However, we still need to take into account the bias in the residuals, defined as the difference between the participant's chronological age and the prediction. As explained in more details under Methods - Biological age definition, we observed a bias in the residuals as a function of chronological age. Participants on the younger end of the chronological age distribution tend to be predicted older than they actually are, whereas participants on the older end of the distribution tend to be predicted younger than they actually are. We need to properly account for this bias when translating a prediction from a more recent instance to an older instance. Let us assume that the average bias in the residuals for participants who are 70 and 80 years old is respectively -2 years and -4 years. After correcting for this bias, the predictions are now respectively 64-(-2)=66 and 78-(-4) = 82. Therefore, the corrected residuals for this participant are respectively -4 years and +2 years, for an average of -1 years. Finally, let us assume that the participant was 60 years old in instance 0. We will assign a single prediction of 60-1=59 years to the participant, but we still need to un-correct for the bias in residuals. Let us assume that the average bias for the residuals at age 60 is +5 years. We will assign a final prediction for the participant of 59+5=64 years.

A key point we would like to highlight here is that we did not actually correct for the bias in the residuals at this step of the pipeline. Instead, we corrected then un-corrected the predictions that we translated from different instances to the instance 0. The actual correction for the residual biases takes place when defining the biological age phenotypes (see Methods - Biological age definition).

To distinguish between raw predictions on the instance 0, and the average predictions reported to the instance 0, we created a new instance which we named instance "*". We refer to these predictions as "participants predictions", as opposed to "samples predictions".

#### Interpretability of the predictions

For elastic nets, we interpreted the models using the values of the regression coefficients. Large absolute values for these coefficients means they played an important role when generating the predictions. For gradient boosted machines we used the feature importances, which are based on the number of times a tree selected each of the variables. Variables with high feature importances were selected more often and are therefore likely to play a key role in predicting chronological age. For neural networks, we estimated the importance of each feature by permuting it randomly between samples before computing the performance of the model. The score of each feature is the difference between the R-Squared value before and after the random permutations. Features whose random permutation leads to a large decrease in the model's performance are estimated to be important predictors of chronological age.

We estimated the concordance between the three different algorithms by computing the Pearson and the Spearman correlations between their feature importances.

#### Non-genetic correlates of accelerated aging

Unlike DNA, biomarkers, phenotypes, diseases, family history, environmental variables and socioeconomics can change over life. As a consequence, we compared each biomarker, phenotype and environmental variable with the accelerated aging of the participant at the time the exposure was measured and we used the "Samples predictions", as opposed to the "Participants predictions" that we used for the identification of genetic correlates (see Methods - Models ensembling - Generating average predictions for each participant).

#### Imputation of the non-genetic X-variables

Most X-variables were not collected on all four instances. Additionally, no X-variables were collected at the same time as the accelerometer data was collected. To identify the non-genetic correlates of accelerated aging, we had to impute the values of the X-variables for the ages of the participants for which they were not available. We considered two imputation methods, which we refer to as the "cross-sectional" and the "longitudinal" imputations.

For the cross-sectional imputation, we computed a linear regression for each X variable as a function of age, adjusting for sex. We then used the slope of the linear regression to extrapolate the value of the XWAS variable at different ages.

For the longitudinal imputation, we first selected, for each X variable, all the participants that had at least two measures taken for this X variable. We then performed a linear regression for each participant. We then averaged the slope of the linear regressions over all the participants of the same sex. Finally, we used this slope to extrapolate the value of the XWAS variable at different ages for all participants depending on their sex, in the same way we did it for the cross-sectional imputation.

It is important to notice that for both the cross-sectional imputation and the longitudinal imputation, data can only be imputed when the XWAS variable has been measured at least once for the participant. This raw measure is then used to extrapolate which value the X variable was likely taking a couple years earlier and/or later.

The advantage of the cross-sectional imputation is larger sample sizes. The advantage of the longitudinal method is that it corrects for generational effects. For example, old people have shorter legs than young people on average [24]. This is not because human legs shrink as we grow older. Instead, people who are old today already had shorter legs when they were young. If the cross-sectional regression is used to impute the length of the participants on instances where it was not measured, it will spuriously assign smaller values to the older samples. In contrast, the longitudinal regression learns the regression coefficient by comparing each participant to themselves as they age and will therefore not capture the generational effect. When used to predict the participants legs' length, it will impute constant values over time. To evaluate which of the two imputation methods should be preferred, we used them to predict X-variables for which we knew the actual values and computed the R-Squared values associated with the predictions. We found that, even with sample sizes as small as 200 samples, longitudinal imputation outperformed cross-sectional imputation. We therefore used longitudinal imputation.

#### X-Wide Association Studies

First, we tested for associations in an univariate context by computing the partial correlation between each X-variable and aging dimensions. To compute the partial correlation between an X-variable and an aging, we followed a three steps process. (1) We ran a linear regression on each of the two variables, using age, sex and ethnicity as predictors. (2) We computed the residuals for the two variables. (3) We computed the correlation between the two residuals and the associated p-value if their intersection had a sample size of at least ten samples. We used a threshold for significance of 0.05 and corrected the p-values for multiple testing using the

Bonferroni correction. We plotted the results using a volcano plot. We refer to this pipeline as an X-Wide Association study [XWAS].

In the supplementary tables and the results, we rank the X-variables subcategories by decreasing percentage of variables associated with accelerated aging (note that the ranking is therefore biased towards categories with fewer variables). For each subcategory, we list the three most associated variables, based on the absolute value of the correlation coefficient. For the exhaustive list, please refer to https://www.multidimensionality-of-aging.net/xwas/univariate_associations.

#### Prediction of accelerated aging

We leveraged the pipeline we built to predict chronological age as a function of scalar biomarkers to predict accelerated auditory aging as a function of the biomarkers, clinical phenotypes, diseases, family history, environmental and socioeconomic variables. We leveraged the same pipeline to identify which features were driving the predictions. We built a model for each X-variables subcategory (Table S3, Table S6, Table S9, Table S12, Table S13, Table S16).

### Supplementary Figures

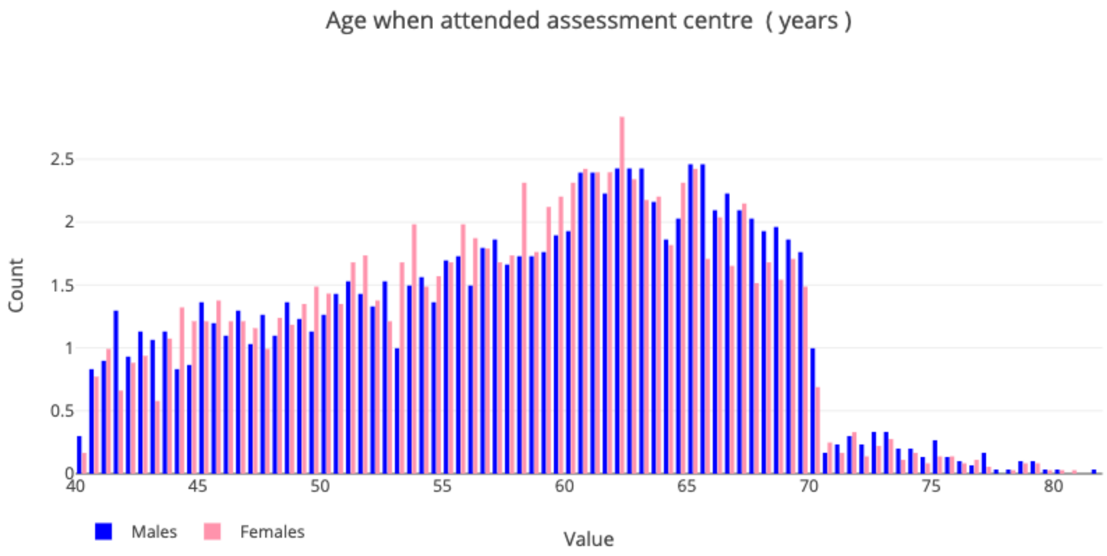


**Figure S1: Demographics of the UK Biobank cohort**

### Supplementary Tables

**Table S1: Feature importances for the models predicting age**

See supplementary data

**Table S2: Pearson and Spearman correlations between the feature importances for different models**

Pearson: Upper right. Spearman: Lower left.

| | Correlation | Elastic Net | GBM | Neural Network |
|---|---|---|---|---|
| Correlation | NA | -0.217 | 0.181 | 0.079 |
| Elastic Net | -0.056 | NA | -0.040 | 0.245 |
| GBM | 0.531 | 0.300 | NA | 0.559 |
| Neural Network | 0.253 | 0.500 | 0.629 | NA |

**Table S3**: **List of biomarkers by subcategories for the Biomarkers Wide Association Study [BWAS]**

See supplementary data

**Table S4: Biomarkers most associated with accelerated auditory aging**

See supplementary data

**Table S5: Biomarkers most associated with decelerated auditory aging**

See supplementary data

**Table S6: List of clinical phenotypes by subcategories for the Clinical Phenotypes Wide Association Study [CWAS]**

See supplementary data

**Table S7: Clinical phenotypes most associated with accelerated auditory aging**

See supplementary data

**Table S8: Clinical phenotypes most associated with decelerated auditory aging**

See supplementary data

**Table S9: List of diseases by subcategories for the Diseases Wide Association Study [DWAS]**

See supplementary data

**Table S10: Diseases most associated with accelerated auditory aging**

See supplementary data

**Table S11: Diseases most associated with decelerated auditory aging**

See supplementary data

**Table S12: List of family history variables by subcategories for the Family History Phenotypes Wide Association Study [FWAS]**

See supplementary data

**Table S13: List of environmental variables by subcategories for the Environmental Wide Association Study [EWAS]**

See supplementary data

**Table S14: Environmental variables most associated with accelerated auditory aging**

See supplementary data

**Table S15: Environmental variables most associated with decelerated auditory aging**

See supplementary data

**Table S16: List of socioeconomic variables by subcategories for the Socioeconomics Wide Association Study [SWAS]**

See supplementary data

**Table S17: Socioeconomic variables most associated with accelerated auditory aging**

See supplementary data

**Table S18: Socioeconomic variables most associated with decelerated auditory aging**

See supplementary data

**Table S19: Hyperparameter space for scalar features-based models Bayesian optimization**

| Algorithm | Hyperparameter | Scale | Low | High |
|---|---|---|---|---|
| Elastic net | alpha | loguniform | -10 | 0 |
| | l1_ratio | uniform | 0 | 1 |

| | | | | |
|---|---|---|---|---|
| Gradient Boosted Machine | num_leaves | quniform | 5 | 45 |
| | min_child_samples | quniform | 100 | 500 |
| | min_child_weight | loguniform | -5 | 4 |
| | subsample | uniform | 0.2 | 0.8 |
| | colsample | uniform | 0.4 | 0.6 |
| | reg_alpha | loguniform | -2 | 2 |
| | reg_lambda | loguniform | -2 | 2 |
| | n_estimators | quniform | 150 | 450 |
| Neural network | learning_rate_init | loguniform | -5 | -1 |
| | apha | loguniform | -6 | 3 |

**Table S20: Nested Cross-Validation pipeline**

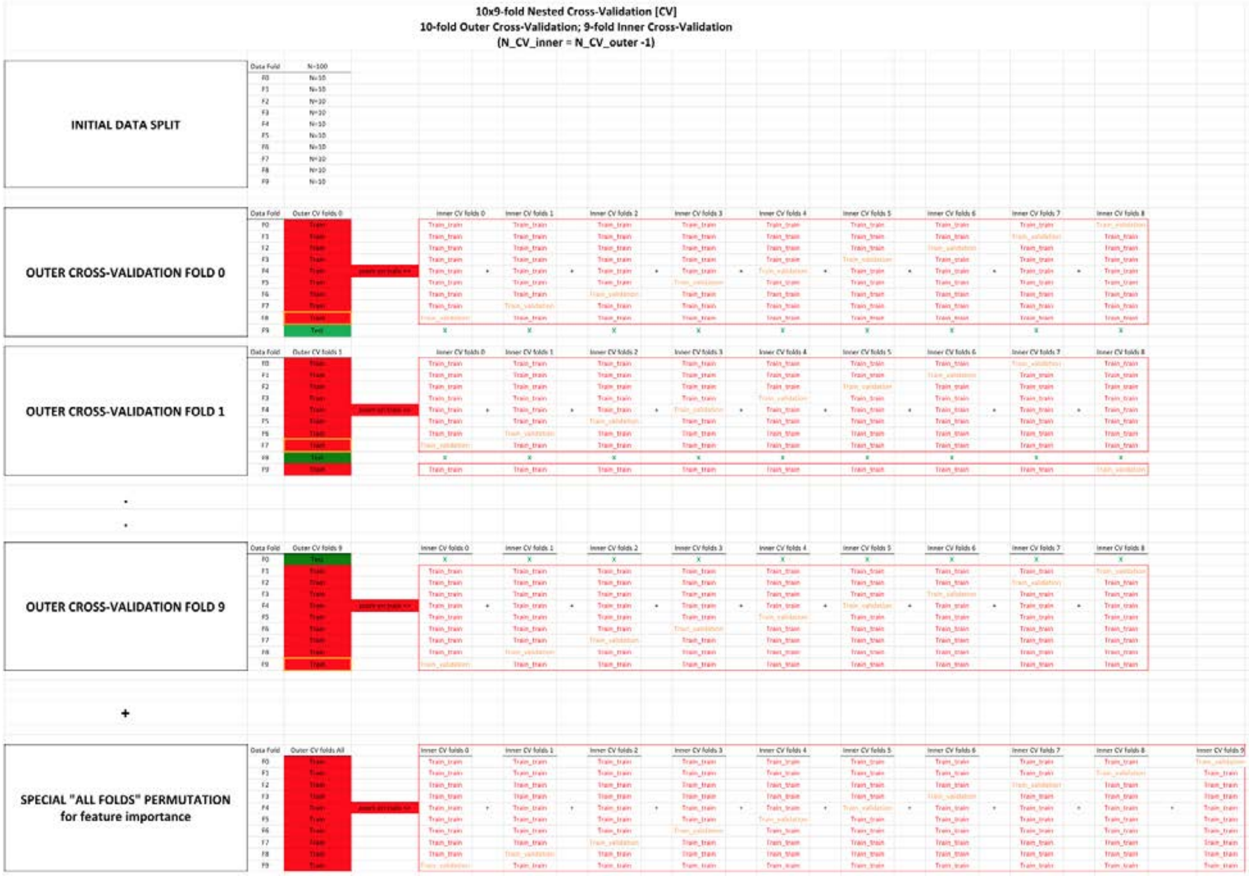


| METHODS: | |
|---|---|
| **General comments** | The Nested Cross-Validation is a normal double split train/val/test method, for which not just one, but the two splits are replaced with a CV instead. |
| **Outer Cross-Validation** | The Outer Cross-Validation is used to replace the split between train + val on one side, vs. test on the other side. Thanks to the Outer Cross-Validation, an unbiased testing prediction can be obtained for every sample.<br>Three common sources of confusion clarified:<br>**Yes,** using an Outer Cross-Validation does mean that the predictions do not all come from the same model. (For exemple for testing, the prediction for each fold comes from a different model.)<br>**No**, the Outer Cross-Validation has ***nothing*** to do with hyperparameter tuning or model selection.<br>Tuning a model only requires one Outer Cross-Validation. In contrast, it requires N_CV_folds distinct Inner Cross-Validations, one for each fold of the Outer Cross Validation. (see below) |
| **Inner Cross-Validation** | The Inner Cross-Validation is used to replace the split between the typical training set and validation set.<br>Thanks to the Inner Cross-Validation, hyperparameter selection can be decided based on a performance calculated on the full training set and validation set sample size, instead of the sole validation set sample size.<br>Two common sources of confusion clarified:<br>#1 - The whole purpose of performing an Inner Cross-Validation is ***hyperparameter selection***.<br>The Inner Cross-Validation tunes the model that will then be used to generate predictions on the left out testing fold.<br>Therefore, the Inner Cross-Validation does ***not*** generate a testing prediction for more samples. That is the role of the Outer Cross-Validation.<br>One last time, the Inner Cross-Validation is ***only*** about improving the hyperparameter tuning of the model.<br>#2 - Tuning a model requires N_CV_folds distinct Inner Cross-Validations, one for each fold of the Outer Cross Validation. In contrast, it only requires one Outer Cross-Validation. (see above) |
| **Special Fold 0** | Special "All folds" permutation is only here for feature importance purposes.<br>We could extract the features importance from the 10 models used to generate the predictions on the testing set, but we would have to take the average of features importances between several model.<br>Addtionally each of these models only saw 90% of the data (10% was left aside for testing purposes).<br>If we only care about feature importance, we actually do not need to leave samples aside for testing. We can use 100% of the data to train a model using a 10-fold (not 9-fold this time, since the test fold can be used too!) ***Inner*** Cross-Validation to train a single model.<br>This model will never be tested, but it does not matter: its role is to provide the best estimate for the feature importance, which it does well.<br>Two common sources of confusion clarified:<br>#1 - The Nested Cross-Validation pipeline and the Special "All Folds" permutation pipeline are ***not*** intertwined and serve different purposes.<br>The purpose of the Nested Cross-Validation pipeline is to generate accurate and a unbiased testing prediction for every sample, whereas the purpose of the Special "All Folds" pipeline is to generate feature importance.<br>#2 - **Yes**, it does mean that the feature importance reported do not correspond to any of the 10 models (trained in the 10 Outer Cross-Validation folds) that generated the final testing predictions.<br>The feature importance comes from a model that was never used to generate predictions. |

| RESULTS: | **Example**: cells corresponding to the values used for each prediction set | **Explanation**: |
|---|---|---|
| **Training** | "E17+G17+I17+K17+M17+O17+Q17+S17+<br>E29+G29+I29+K29+M29+O29+Q29+U29+..." | To obtain the training prediction for the Data Fold F1, take the mean of every training prediction available on this data fold (10*8=80 of them). |
| **Validation** | "E25" | To obtain the validation prediction for the Data Fold F1, take the prediction outputted by the model trained on the remaining 8 data folds with the best hyperparameters values obtained for this Outer Cross-Validation fold.<br>These validation predictions can then be used to tune the ensemble models. /!\ Warning /!\: This can only be properly done if data folds split used for the Outer Cross-Validation and the Inner Cross-Validation is the same. |
| **Testing** | "C26" | No extra step is needed for the testing prediction for the Data Fold 1. Simply take the only testing prediction available for this fold. |
| **Feature importance:** | N/A | Extract the features importance from the special "outer fold" "All".<br>If want to compute the standard deviation for the feature importance as well, we can leverage the feature importance computed on each of the Outer Cross-Validation folds and take the square root of their variance. |

SUPPLEMENTARY:

**Comparison between Nested Cross-Validation and other validation methods:**

**#1**-Single split train/test.
Split train/test works when there are no hyperparameters to tune. If one uses a simple train/test split and tune the hyperparameters on the testing set, some overfitting is to be expected.

**#2**-Double split train/validation/test.
Because of the above, a train/validation/test double split is commonly used when hyperparameters tuning is required.
The training is performed on the training set, the tuning of the hyperparameters on the validation set, and the evaluation of the unbiased model performanc on the untouched test dataset.

**#3**-Cross-Validation(train/val) and split (train+val)/test.
With the pipeline #2 (double split train/validation/test), the tuning of the hyperparameters is not leveraging the dataset as much as with a Cross-Validation, as only one data fold is used to tune the hyperparameters.
Therefore machine learning practitioners usually replace the "inner split" (the training/validation split), by a Cross-Validation instead, but they keep the "outer split" (between the training+validation dataset used for their Cross-Validation and the testing set).

**#4**-Split train/val and Cross-Validation((train+val)/test).
Pipeline #3 only generate an unbiased testing prediction for a fraction of the dataset (the testing set).
To generate an unbiased prediction on every sample of the dataset, pipeline #4 can be used.
However, because there is a simple split between the training set and the validation set (one for each Cross-Validation fold), that means that the hypperparameter tuning only leverages a fraction of the dataset.
Pipeline #4 is preferred when generating an unbiased testing prediction for every sample is needed but the models are too time consuming to train to be able to afford a Nested Cross-Validation (#5).
Pipeline #4 is therefore the pipeline we used to predict chronological age using medical images or videos, for example.

**#5**-Nested Cross-Validation.
To fully leverage the whole dataset, a Nested Cross-Validation replaces both the outer split (train+val/test) and the inner splits (train/val) by a Cross-Validation, hence the nesting.
It requires the training of a large number of models, and it is therefore usually reserved for models that can be trained relatively quickly.
In our case, we used it to train our scalar features-based models (elastic nets, light gradient boosted machines, and shallow fully connected neural networks).
